# Aging and COVID-19 mortality: A demographic perspective

**DOI:** 10.1101/2020.10.15.20213454

**Authors:** Isaac Sasson

**Affiliations:** Department of Sociology and Anthropology, The Herczeg Institute on Aging, Tel Aviv University

## Abstract

Demographers have emphasized the importance of age in explaining the spread and impact on mortality of COVID-19. However, the relationship between COVID-19 with age should be contextualized in relation to other causes of death. This study set out to compare the age pattern of COVID-19 mortality with other major causes of death and across countries, and to use these regularities to impute age-specific death counts in countries with partial data. Using US vital Statistics, the COVID-19 mortality doubling time in a Gompertz context was compared with 65 major causes of death. COVID-19 fatality and mortality doubling times were similarly compared within and between 27 countries, using harmonized demographic databases of confirmed COVID-19 cases and deaths. Several findings are supported by the empirical evidence. First, COVID-19 mortality increases exponentially with age at a Gompertz rate near the median of aging-related causes of death, as well as pneumonia and influenza. Second, COVID-19 mortality levels are 2.8 to 8.2 times higher than pneumonia and influenza across the adult age range. Third, the relationship between both COVID-19 fatality and mortality with age varies considerably across countries. Fourth, COVID-19 deaths by age are imputed for Israel as a case in point. In conclusion, the increase in COVID-19 mortality with age resembles the population rate of aging. Country differences in the age pattern of COVID-19 fatality and mortality may point to differences in underlying population health, standards of clinical care, and data quality. This study underscores the need to contextualize the age pattern of COVID-19 mortality in relation to other causes of death.

## 1. Introduction

There is mounting clinical evidence that the risk of dying from Coronavirus disease 2019 (COVID-19) is associated with age (e.g., Zhou et al., 2020). Clinicians noted early on that COVID-19 fatality rises steadily with age; unlike other respiratory diseases, it did not present the typical U-shaped curve of heightened risk among both infants and older adults (Raoult et al., 2020). The World Health Organization (2020) and the US Centers for Disease Control and Prevention (2020a) have both defined older adults—individuals aged 60 and 65 and older, respectively—as a vulnerable group. In response, as the COVID-19 pandemic broke out, governments worldwide recommended or instructed older adults to remain in social isolation for a period lasting weeks or months (Brooke & Jackson, 2020). These simplified accounts of the relationship between COVID-19 and age may have contributed to the portrayal of COVID-19 as a “disease of the elderly,” in the US and elsewhere, which some have argued was a key factor in the failure to contain the outbreak (Germani et al., 2020; Nowroozpoor et al., 2020). Nearly one year into the pandemic, there is now sufficient demographic data to establish key facts about the relationship between age and COVID-19 mortality.

Demographers were among the first to point out the importance of age in explaining COVID-19’s spread and impact on mortality (e.g., Dowd et al., 2020; Dudel et al., 2020; Kashnitsky & Aburto, 2020). Several studies found that the age pattern of COVID-19 mortality resembles that of all-cause mortality, and similarly follows the Gompertz law of mortality (Goldstein & Lee, 2020; Promislow, 2020). Others have noted considerable variation in the rate in which COVID-19 mortality increases with age across countries (Demombynes, 2020), pointing perhaps to differences in age-specific infection rates, standards of medical care, and the classification of COVID-19 deaths. Yet, there are several outstanding questions about the relationship between age and COVID-19 mortality:

1. How does the age pattern of COVID-19 mortality compare with other major causes of death?
2. How does COVID-19 mortality compare with pneumonia and influenza (P&I), with respect to both absolute levels and proportional increase with age?
3. How do the age patterns of COVID-19 mortality and fatality vary across OECD and high-income countries?
4. How can these regularities be used for imputing COVID-19 deaths by age in countries with partial data?

Building upon prior research, this study analyzed COVID-19 mortality and fatality data from the US and 26 other OECD and high-income countries. Several findings are supported by the empirical evidence. First, consistent with previous studies, COVID-19 mortality and fatality follow the Gompertz law in most countries. Second, COVID-19 mortality increases with age at a rate similar to the median aging-related cause of death, as well as annual P&I mortality. However, COVID-19 mortality in the US is higher than average P&I mortality by a factor of 2.8 to 8.2 across the adult age range. Third, the relationship between age and both COVID-19 mortality and fatality varies considerably across OECD and high-income countries. Lastly, a simple method for imputing COVID-19 death counts by age is developed and applied to Israel as a case in point.

## 2. Data and methods

Already in 1825, Benjamin Gompertz noted that beyond early adulthood all-cause mortality rises exponentially with age, taking the form (Gompertz, 1825):

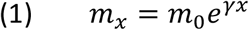

where *m_x_* is the mortality rate at age *x* and *m*_0_ is the baseline mortality rate of young adults. The exponential rate of increase in mortality rate with age, *γ*, has often been equated with the population rate of aging (Ricklefs & Scheuerlein, 2002). The rate of aging is closely linked to the mortality rate doubling time (MRDT), the average number of years in which the mortality rate doubles:

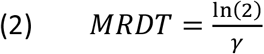

While the interpretation of the Gompertz slope as the rate of aging has been criticized for conceptual reasons (de Grey, 2005), the Gompertz hazard function remains a useful model for senescent mortality (Gavrilov & Gavrilova, 2019) as well as for the relationship of specific causes of death with age (Juckett & Rosenberg, 1993). A larger Gompertz slope coefficient implies, if the model fits well, a steeper progression of mortality with age. Conversely, causes of death that progress rapidly with age will have *low* mortality rate doubling time, whereas those that progress slowly with age will have *high* doubling time.

The analysis consisted of several steps. First, the MRDT between ages 25 and 84 was estimated for 113 underlying causes of death using US vital statistics data from 2018 (Centers for Disease Control and Prevention, 2020b). Only causes of death for which the Gompertz model was a good fit (R^2^ > 0.95) were retained in the analysis. The resulting 65 causes of death were compared with COVID-19 mortality doubling time, estimated from provisional US deaths counts (National Center for Health Statistics, 2020b).

Second, COVID-19 age-specific mortality rates were compared with those of P&I between 1999 and 2018. Adjusting the exposure time is critical when comparing mortality rates because COVID-19 is an ongoing pandemic (Heuveline & Tzen, 2020). Annual P&I mortality rates were therefore adjusted to the same calendar period as COVID-19 provisional death counts—epidemiological weeks 6 through 47—using historical data from the National Center for Health Statistics (2020b).

Since COVID-19 mortality might vary by geographic regions, viral strain, or other factors (Brufsky, 2020; Wilson et al., 2020), it is important to establish whether the US findings can be generalized to other countries. Thus, the third step was to compare the relationship between age and COVID-19 fatality (and mortality, where possible) across different countries. Age-specific case fatality rates—i.e., the number of deaths per confirmed COVID-19 cases in each age group—were estimated using COVerAGE-DB, a harmonized cross-national database of confirmed COVID-19 cases and deaths by age (Riffe et al., 2020). At the time of analysis, the database had complete COVID-19 cases and death counts by age for 33 OECD and high-income countries, as currently defined by the World Bank (2020). For a small set of those countries, it was also possible to estimate COVID-19 mortality rates by age using the French National Institute for Demographic Studies COVID-19 demographic database (INED, 2020). Much like the MRDT, the case-fatality rate doubling time (FRDT) was estimated using Equation 1, substituting fatality rates for mortality. Although the two measures are not directly comparable, their relationship with age will be similar if attack (infection) rates do not vary markedly by age. This assumption was evaluated for the US and 9 other countries that were present in both databases. A difference between the FRDT and MRDT could indicate that certain age groups are disproportionally likely to either become infected or to be diagnosed, a phenomenon known as surveillance bias (Haut, 2011).

Lastly, the Gompertz model was used to impute COVID-19 deaths by age when only the total number of deaths and age-specific exposures (diagnosed cases) were known. The cumulative number of COVID-19 deaths by country is widely available (Dong et al., 2020). By contrast, age-specific death counts are far more scarce and often incomplete (Riffe et al., 2020). This was, for example, the case of Israel, where the Ministry of Health publicly released age-specific COVID-19 exposures but suppressed the number of deaths by age due to confidentiality concerns. By borrowing information from other countries, the Gompertz model provides a simple method for imputing the number of COVID-19 deaths by age.

Assuming that COVID-19 fatality rates follow Equation 1, substituting fatality for mortality rates, the number of deaths as a function of age is given by

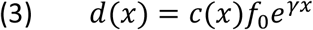

where *c(x)* is the number of diagnosed cases by age and *f_0_* is the baseline fatality rate. The expected number of deaths by age can then be imputed using Equation 3 for an observed total number of deaths, *D*, and a specified slope parameter, 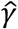, solving for *f_0_*:

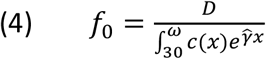

Values for 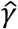 can be borrowed from a reference country or distribution, with *f_0_* serving as a scaling parameter to ensure that deaths summed across age groups match the known total. In other words, the number of deaths by age equal the Gompertz fatality schedule weighted by observed age-specific exposures.

## 3. Results

### 3.1 How does the relationship between COVID-19 mortality and age compare with other causes of death?

The mortality rate doubling time of major cause-of-death categories is shown in Figure 1. Age is a strong predictor of each of those causes because poorly fitting models have been excluded. In 2018, the MRDT varied in the US from 3.2 (90% CI 3.1–3.3) years for Alzheimer’s disease (more age dependent) to 21.1 (90% CI 18.3– 23.8) years for Asthma (less age dependent), with a median of 7.3 years across all 65 cause of death categories, excluding COVID-19. For example, the MRDT was estimated at 5.3 (90% CI 4.7–5.9) years for chronic lower respiratory diseases; 6.6 (90% CI 5.3–7.9) years for acute myocardial infarction; 7.0 (90% CI 6.0–8.0) years for malignant neoplasms; 8.0 (90% CI 7.1–9.0) years for diabetes; and 10.9 (90% CI 9.2– 12.5) years for kidney infections. The COVID-19 MRDT, based on US provisional data through 21 November 2020, was estimated at 7.6 (90% CI 7.2–8.0) years. In other words, it is near the median doubling time for all aging-related causes of death. Moreover, the relationship between COVID-19 mortality and age is not markedly different from that of Pneumonia & Influenza, which in 2018 had an estimated MRDT of 7.7 (90% CI 7.4–8.0) years.

**Figure 1.**
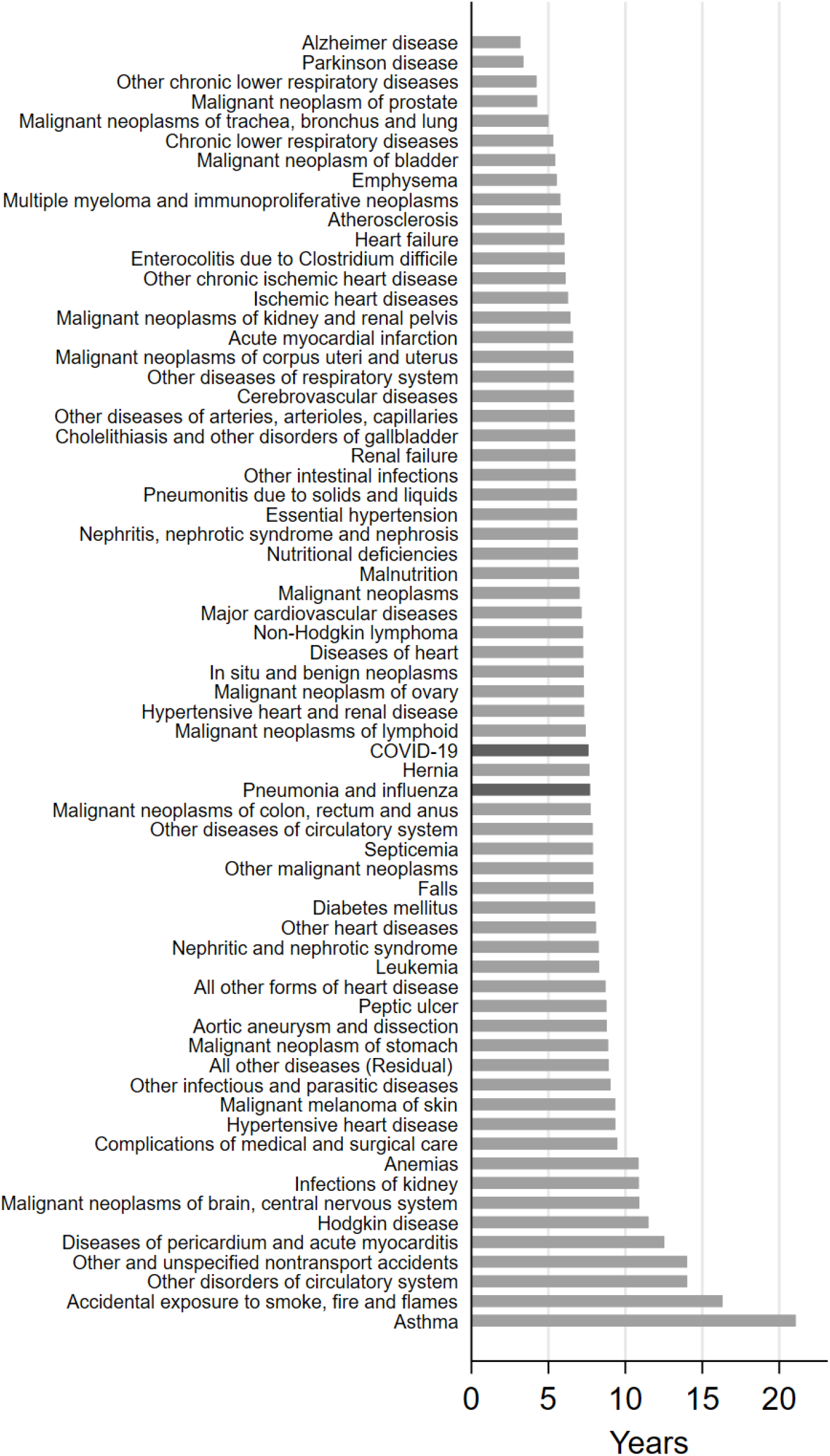
Mortality rate doubling time of COVID-19 and major causes of death in 2018, United States. Note: Mortality rate doubling time estimated from 2018 Multiple Cause of Death data (Centers for Disease Control and Prevention, 2020b); estimate for COVID-19 based on provisional death counts through 21 November 2020 (National Center for Health Statistics, 2020b).

### 3.2 How does COVID-19 mortality compare with Pneumonia & Influenza?

Figure 2 compares COVID-19 age-specific mortality with a 20-year series of P&I mortality. Since COVID-19 is an ongoing pandemic, annual P&I mortality rates have been adjusted to the equivalent exposure period based on historical seasonal data (i.e., the proportion of P&I deaths occurring between epidemiological weeks 6 through 47). Figure 2 reveals that COVID-19 absolute mortality levels are several times higher than average P&I mortality. These stark differences cannot be attributed to sampling error (the margin of error of COVID-19 age-specific mortality, indiscernible on a logarithmic scale, was less than ±6% at any age group and thus not shown in the figure). COVID-19 mortality was 5.2 times greater than average P&I mortality at ages 25-34; 6.6 times greater at 35-44; 8.1 times greater at 45-54; 8.2 times greater at 55-64; 6.5 times greater at 65-74; 4.3 times greater at 75-84; and 2.8 times greater at age 85 and over. However, the proportional rate of increase in COVID-19 mortality with age was well within the inter-annual variation in P&I mortality, and similar to the latter’s multiyear average. From 1999 to 2018, the P&I mortality-rate doubling time averaged 7.4 (90% CI 7.2–7.6) years, ranging from 6.8 (90% CI 5.9–7.8) to 8.9 (90% CI 7.3–10.6) years over the 20-year period. The COVID-19 mortality doubling time was estimated at 7.6 (90% CI 7.2–8.0) years.

**Figure 2.**
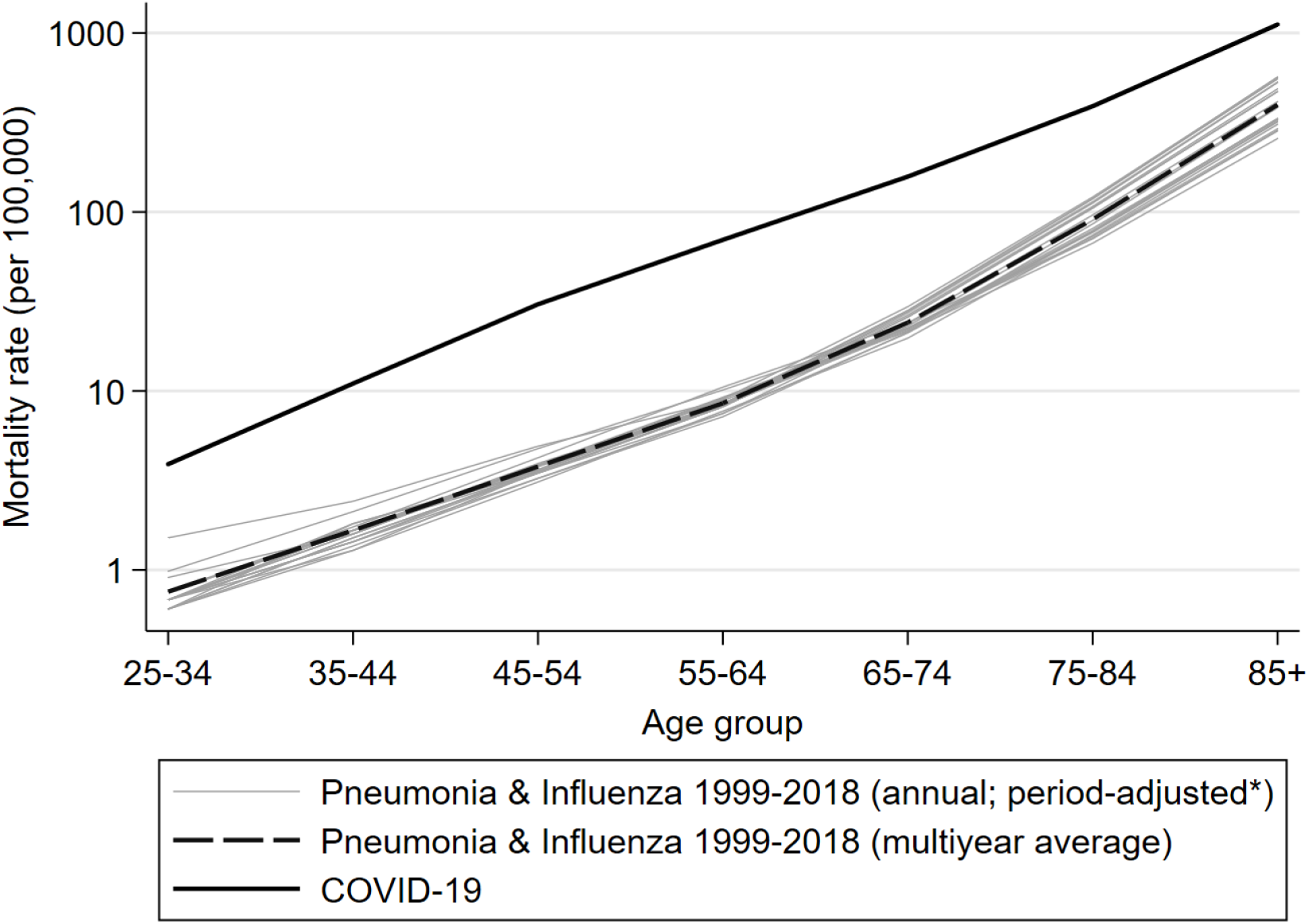
Age-specific mortality rates of COVID-19 (2020) and Pneumonia & Influenza (1999–2018), United States. Note: COVID-19 mortality rates based on provisional US death counts from 1 February to 21 November 2020 (National Center for Health Statistics, 2020b); Pneumonia & Influenza mortality rates based on vital statistics data (Centers for Disease Control and Prevention, 2020b) and adjusted to the same calendar period as COVID-19 provisional data.

### 3.3 How do age patterns of COVID-19 mortality and fatality vary across countries?

The third objective was to determine whether the relationship between age and both COVID-19 fatality and mortality varies across OECD and high-income countries. The Gompertz model fit age-specific fatality rates well (R^2^ > 0.95) in 27 of the 33 countries examined. The FRDT of COVID-19, shown in Figure 3, varied from 4.5 (90% CI 4.1–4.9) years in Denmark to 9.9 (90% CI 8.6–11.2) years in Mexico, with a median FRDT of 5.3 years and a mean of 5.9 years. For 10 of the 27 countries, it was possible to estimate in addition the MRDT of COVID-19 (also shown in Figure 3). MRDT values ranged from 5.1 (90% CI 4.8–5.3) years in Denmark to 7.6 (90% CI 7.2–8.0) years in the US. With few exceptions, estimated FRDT and MRDT values were substantively similar across countries (shown with 90% confidence intervals in Figure 3). In the US and in Germany, however, the MRDT was substantially higher than the FRDT by 0.9 and 1.0 years, respectively. The MRDT in Spain, by contrast, was lower than the FRDT by 1.4 years. The ratio of COVID-19 mortality to fatality rates by age, an indirect measure of COVID-19 attack rates, is further explored in the US context in Figure A1 in the Appendix. The results confirm that the ratio of COVID-19 mortality to fatality indeed tends to decrease with age. Overall, the findings in this section suggest that in the majority of countries both COVID-19 mortality and fatality follow the Gompertz function (while the model pertains to adult mortality, only 0.25% of deaths across the countries examined occurred under age 25). The rate in which COVID-19 mortality and fatality increase with age, however, varies considerably between countries.

**Figure 3.**
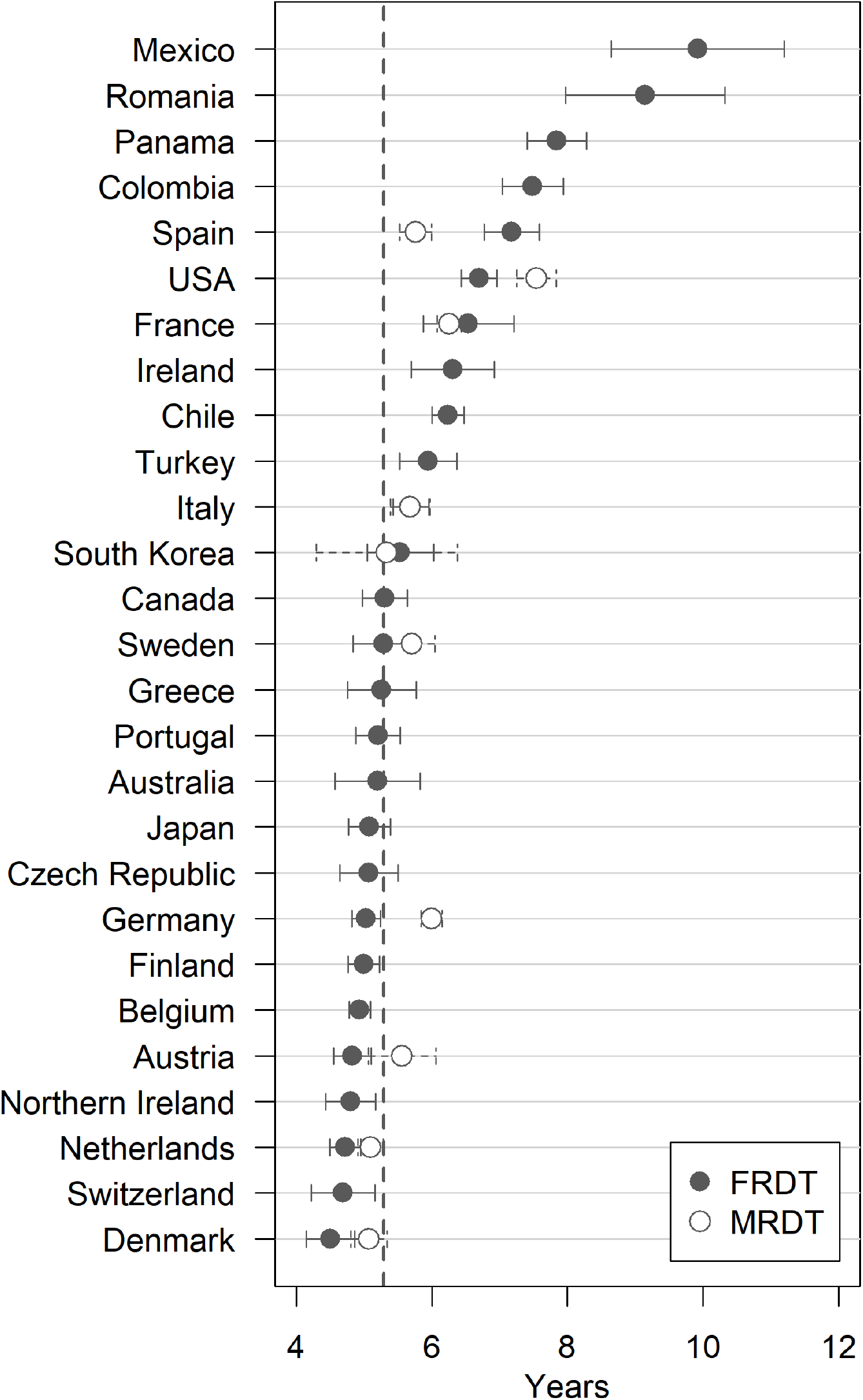
COVID-19 fatality and mortality doubling times across OECD and high-income countries (with 90% confidence intervals). Notes: FRDT = fatality rate doubling time; MRDT = mortality rate doubling time. Median FRDT marked by dashed line. Last COVerAGE-DB reporting date: Australia (25 Nov); Austria (25 Nov); Belgium (20 Nov); Canada (17 Oct); Chile (9 Nov); Colombia (19 Jul); Czech Republic (15 Nov); Denmark (24 Nov); Finland (23 Nov); France (26 May); Germany (24 Nov); Greece (9 Nov); Ireland (24 May); Italy (20 Oct); Japan (18 Aug); Mexico (1 Aug); Netherlands (24 Nov); Northern Ireland (24 Nov); Panama (4 Jul); Portugal (21 Nov); Romania (15 Apr); South Korea (22 Nov); Spain (21 May); Sweden (23 Nov); Switzerland (24 Nov); Turkey (19 Oct); USA (14 Nov).

### 3.4 Imputing COVID-19 death counts by age

COVID-19 mortality and fatality seem to follow the Gompertz law in most countries. This regularity can be used for imputing the expected number of deaths by age when only the total is known. Israel, as a case in point, is one of the countries in COVerAGE-DB that reported the number of confirmed COVID-19 cases, but not deaths, by age. Nevertheless, the cumulative number of deaths is published daily by the Israel Ministry of Health. A total of 2,754 COVID-19 deaths have been reported through 21 November 2020 (Israel Ministry of Health, 2020); only one of those deaths, according to various media accounts, occurred under age 25. Under the Gompertz model, the expected number of deaths at each age group can be estimated from the number of exposures by age and the total number of deaths (Equations 3 and 4). Values for the Gompertz slope, *γ*, were bootstrapped from the empirical distribution across the 27 countries in Figure 3, incorporating in addition the sampling variability of 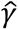 (here assumed to follow, in each country, a scaled 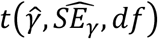 distribution).

Figure 4 shows the expected number of COVID-19 deaths by age in Israel for select Gompertz slope values (10th, 50th, and 90th percentiles). According to the median scenario, for example, 290 deaths (10.5%) are expected under age 65, 1,124 deaths (40.8%) between ages 65 and 84, and 1,339 deaths (48.6%) over age 85. The steeper the Gompertz slope, the more peaked the distribution of deaths at older ages. For the 10th percentile, 27.5% of the deaths occur under age 65 and 28.4% over age 85, whereas for the 90th percentile the respective figures are 6.5% and 56.9%.

**Figure 4.**
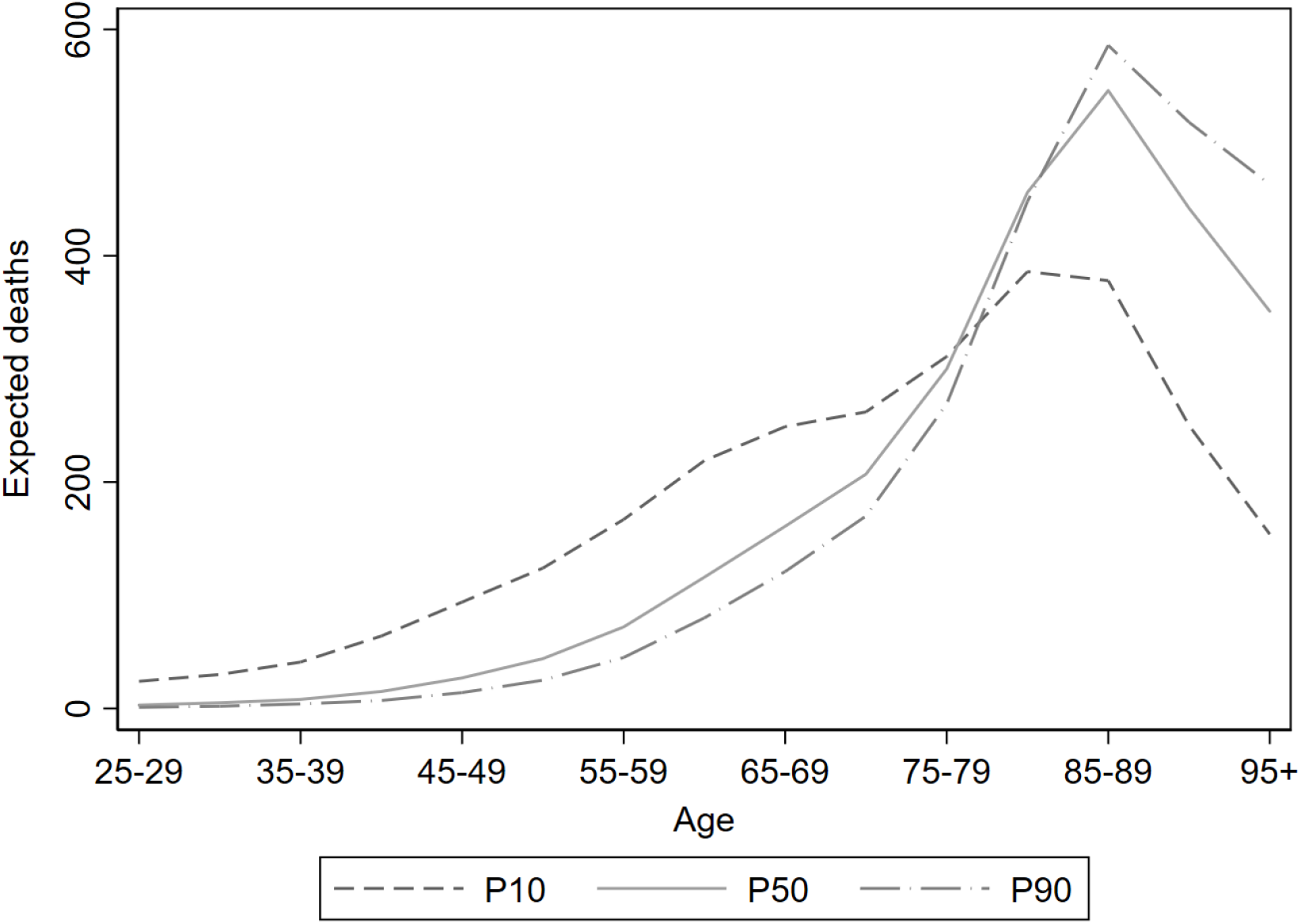
Expected number of COVID-19 deaths by age under three Gompertz slope values, Israel. Note: 10th, 50th, and 90th percentiles of the Gompertz slope distribution were bootstrapped from 27 OECD and high-income countries.

## 4. Discussion

It is hardly news that the risk of mortality increases with age—a relationship that has only strengthened throughout the epidemiologic transition (Salomon & Murray, 2002). COVID-19 mortality is no different, rising exponentially with age. That COVID-19 follows the Gompertz law of mortality, however, is in no way a given (Goldstein & Lee, 2020). The Spanish Flu, in sharp contrast, wrought havoc among 20 to 40 year-olds, whose mortality was substantially higher than their elders (Ma et al., 2011). Consistent with prior research, the present study finds that COVID-19’s relationship with age is approximately log-linear over age 25 and its slope sits near the median of aging-related causes of death. Furthermore, the relationship between age and COVID-19 mortality resembles that of annual pneumonia and influenza mortality over the past 20 years. COVID-19 mortality levels in the US, however, are currently 2.8 to 8.2 times *higher* than average pneumonia and influenza mortality across the adult age range.

Taking a comparative approach, the rates in which COVID-19 mortality and fatality increase with age vary considerably across OECD and high-income countries. While the Gompertz model was a good fit for both COVID-19 mortality and fatality in most countries, its slope varied from one country to the next. In Mexico, for example, the fatality rate doubling time was nearly twice as high as in Denmark. Differences in the age pattern of COVID-19 fatality and mortality between countries may reflect differences in the prevalence of risk factors, standards of clinical care, readiness of healthcare systems, or data quality and reporting practices of COVID-19 deaths (Carinci, 2020; Pasquariello & Stranges, 2020).

In 10 of the 27 countries examined, it was also possible to compare how COVID-19 mortality rates increase with age. In five of those countries, the MRDT and FRDT differed by more than 0.5 years in absolute value. Spain in particular stood out with an estimated MRDT that was 1.4 years *lower* than its FRDT. However, Spain’s data were last reported in May 2020 and may be less reliable, because case-fatality estimates are dynamic and tend to peak in the early stages of the pandemic, before leveling off (Ghayda et al., 2020). In the US, on the other hand, COVID-19 MRDT was *higher* than FRDT—7.6 (90% CI 7.2–8.0) years compared with 6.7 (90% CI 6.4–7.0) years, respectively. In other words, COVID-19 mortality in the US tended to rise less sharply with age relative to fatality. This discrepancy suggests a decline in COVID-19 attack rates with age, consistent, for example, with older adults taking greater precautions against COVID-19.

Although chronological age is one of the most widely available demographic attributes, it is important to note that biological aging mechanisms are more predictive of COVID-19 mortality (Mueller et al., 2020). Individuals age at different rates and chronological age often masks substantial variation in frailty (Mitnitski et al., 2002). Treating COVID-19 as a “disease of the elderly” may foster ageist or paternalistic views and fuel intergenerational tensions against the backdrop of a global health crisis (Ayalon et al., 2020). In spite of the strong relationship between COVID-19 mortality and chronological age, resembling other aging-related causes of death, there is no natural age cutoff to distinguish between low and high risk of COVID-19 mortality. Any such cutoff would be invariably arbitrary and depend on societal norms regarding accepted levels of risk (Glynn, 2020). The findings in this study underscore the need to contextualize COVID-19 in relation to other causes of death with respect to both mortality levels and the relationship with age.

## Data Availability

All data are publicly available.

https://wonder.cdc.gov/

https://osf.io/mpwjq/

https://dc-covid.site.ined.fr/en/

## Acknowledgments

The author thanks Alex Weinreb, Alyson van Raalte, Liat Ayalon, and My Hedlin for helpful comments.

## Appendix

**Figure A1.**
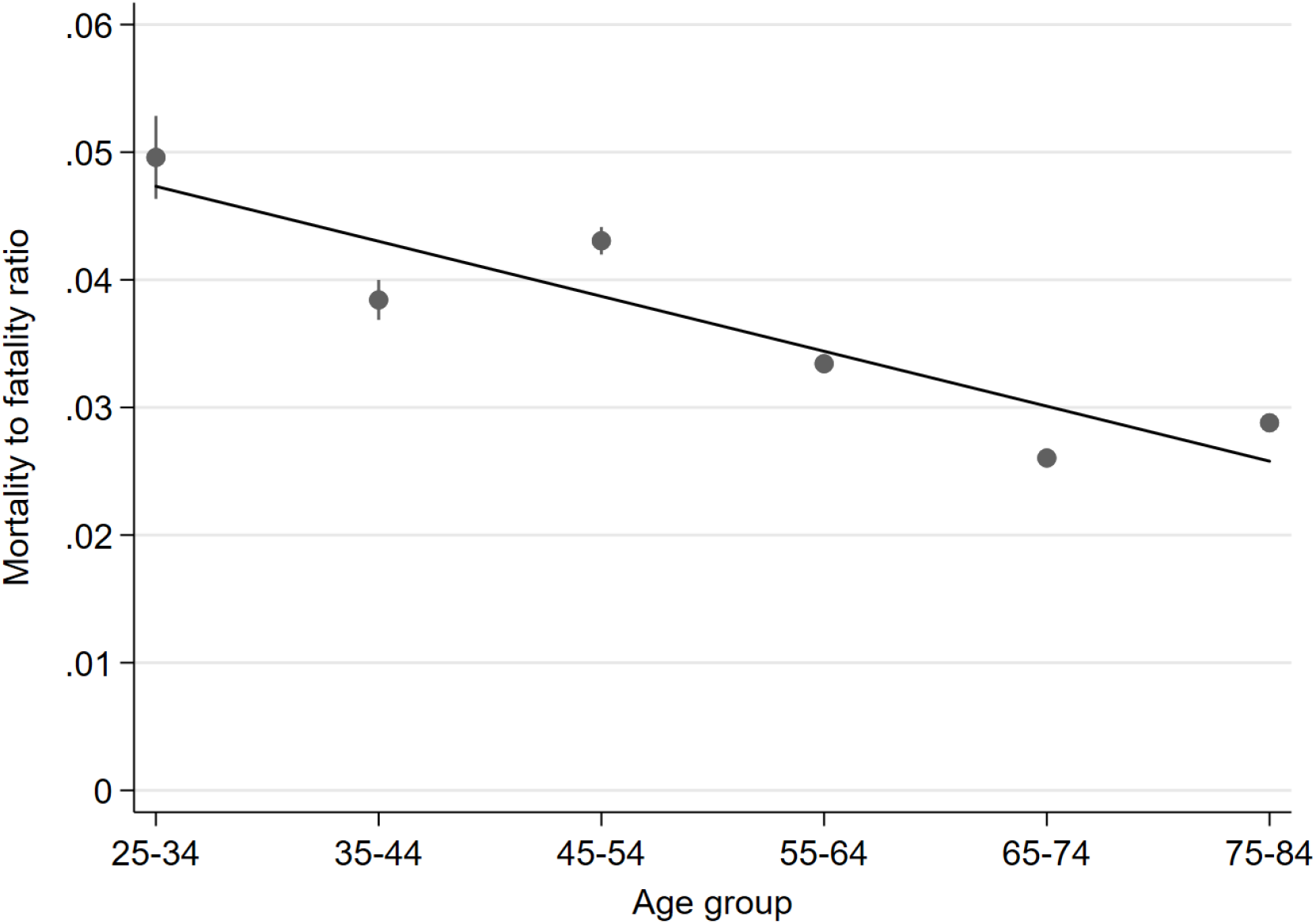
Ratio of COVID-19 mortality to fatality rate by age, United States. Note: Point estimates shown with 95% confidence intervals. Linear fit marked by solid black line.

